# Adolescent autism and autoimmune diagnoses linked to infant gut bacteria whose prevalence is associated with at-risk genetics and/or diet

**DOI:** 10.1101/2021.06.07.21258510

**Authors:** Patricia L. Turpin, Angelica P. Ahrens, Jordan T. Russell, Erik Kindgren, Meghan A. Berryman, Jorma Ilonen, Mark A. Atkinson, Desmond A. Schatz, Eric W. Triplett, Johnny Ludvigsson

**Affiliations:** Department of Microbiology and Cell Science, University of Florida, Gainesville, FL, 32611, USA; Department of Psychiatry, University of Connecticut School of Medicine, Farmington, CT, 06030, USA; Skaraborgs Hospital Skövde, 541 85 Skövde, Sweden; Immunogenetics Laboratory, Institute of Biomedicine, University of Turku, FI-20014 Turku, Finland; Department of Pathology, University of Florida Diabetes Institute, Gainesville, 32610, FL, USA; Department of Pediatrics, College of Medicine, University of Florida, Gainesville, 32610, FL, USA; Crown Princess Victoria’s Children’s Hospital, Region Östergötland, Division of Pediatrics, Linköping University, Linköping, SE 58185, Sweden

## Abstract

The earliest predictors of future autoimmune diseases are a series of autoantibodies that are rarely evaluated and very within and between diseases. In addition, autoantibodies often appear just prior to disease onset. All of these factors make it difficult to apply interventions that might prevent disease. Earlier predictors of disease are needed. Here, a general population cohort was used to assess whether gut bacterial biomarkers could be identified prior to disease. Gut microbiome analysis on 1741 one-year old Swedish children was performed on samples collected in the late 1990s. These children were then followed for 18 years for the incidence of five autoimmune diseases and autism. Specific bacterial strains in the gut microbiome of one-year-old children have been identified as exclusive to the 96 subjects (cases) who acquired type 1 diabetes, celiac disease, hypothyroidism, Crohn’s disease, juvenile idiopathic arthritis, or autism over their next 18 years. None of these strains were found in the 1645 children (controls) who did not acquire any of these diseases. Ten other strains were exclusive to those who remained disease-free. In most cases, the presence or absence of these bacteria were strongly associated with: 1) high-risk class II human leukocyte antigen (HLA) alleles; 2) dietary factors; or 3) a combination of HLA genetics and diet. These results have three significant implications: 1) certain class II HLA haplotypes may serve as bacterial gatekeepers early in life, altering microbiome composition thereby creating the potential for dysbiosis and inflammation; 2) the gut microbiome dysbiosis and inflammation during infancy, largely derived from host HLA genetics and diet, may be a common precedent to all five autoimmune diseases and autism; and 3) HLA gatekeeping may prevent gut colonization of beneficial bacteria in those genetically at-risk individuals who could most benefit from probiotic therapy.

**Funding:** JDRF, Swedish Child Diabetes Foundation, Swedish Council for Working Life and Social Research, Medical Research Council of Southeast Sweden, Regions Östergötland, and Linköping University.

## Main text

Autoimmune disease susceptibility is strongly linked to class II human leukocyte antigen (HLA) allele combinations.^1^ However, autoimmune disorder incidence continues to increase in many developed countries—a finding that cannot be explained by genetic risk alone.^2^ Gut microbiome associations with human autoimmunity have been described for several autoimmune conditions including type 1 diabetes,^3-7^ celiac disease,^8^ juvenile idiopathic arthritis,^9,10^ Crohn’s disease,^11^ and hypothyroidism.^12^

Heritability of autism may be as high as 50-90% and correlation to specific HLA genotypes has been observed. ^13-16^ However, as no autoantibodies for this disorder have been discovered to date, autism is not considered an autoimmune disease. Nevertheless, the gut microbiome is influenced by HLA in those at genetic risk for type 1 diabetes, ankylosing spondylitis, and rheumatoid arthritis and the gut microbiome does differ in those subjects with autism spectrum disorders compared to controls.^17-19^

Most large cohort studies designed to determine the triggers of autoimmune disease include only children at high genetic risk for autoimmunity based on HLA genotype. As a result, these studies cannot assess the influence of genetics over other triggers of disease onset and fail to identify the roles of genetic-environmental interactions in autoimmunity development.^17^ Here stool samples from the general population cohort, All Babies in Southeast Sweden (ABIS), were used to identify gut microbiome markers at one year of age that are associated with disease onset. Currently, autoantibodies are the earliest clinical biomarkers used to predict eventual autoimmune disease cases. When determined, autoantibody markers are subject to infrequent evaluation, thereby reducing their value as predictors of disease. Also, autoantibody markers vary depending on the autoimmune disease in question and can even vary within a disease, such as in type 1 diabetes.^20^

Our initial discovery on the influence of HLA genetics on the gut microbiome at one year of age was conducted in the general population cohort called All Babies in Southeast Sweden (ABIS).^17^ This cohort has data on health and disease of ABIS subjects up to the age of 19. For this effort, the microbiome of stool samples was analyzed from 1,741 ABIS one-year-old children with disease outcomes of type 1 diabetes, celiac disease, juvenile idiopathic arthritis, Crohn’s disease, hypothyroidism, and autism by age 19 years. In that process, a series of gut microbiome biomarkers that were significantly associated with chronic disease, even though they were diagnosed at an average of 13.5 years after stool collection. ABIS cohort data also allowed us to assess whether these biomarkers were associated with specific HLA alleles, diet, or both.

### Cohort Description

The ABIS general population cohort was designed to identify environmental and genetic factors associated with autoimmune diseases.^21^ This study includes 1,741 ABIS subjects at one year of age. Microbial composition was analyzed in all 1,741 one-year-old stool samples collected. Sequencing the V3-V4 region of the 16S rRNA gene resulted in 15,519 amplicon sequencing variants (ASVs) and 331 genera. Presence of HLA risk alleles were determined from the available blood spots of 1,718 children. Dietary data collected included breastfeeding (introduction, exclusivity, and total months), fruit, vegetables, eggs, meat, gluten introduction, and other categories. Other data included gestational age at birth, mode of delivery, number of siblings and pets in the household, infectious episodes, antibiotic use during pregnancy, and other factors. Cases were defined as those ABIS subjects who later in life acquired type 1 diabetes, celiac disease, juvenile idiopathic arthritis, Crohn’s disease, hypothyroidism, or autism versus the controls who did not acquire any of these diseases up to 19 years of age. With those definitions, there are 96 cases and 1,645 controls in this study. Mean age of disease diagnosis and gender for the cases is summarized in Table 1.

**Table 1.** ABIS cohort disease outcomes including number, mean age of diagnosis in years and the range of ages in years of disease onset. Four of the children were diagnosed with more than one of these diseases. The numbers below refer only to the first diagnosis of a disease. Prevalence and mean number of 16S rRNA copies per g of stool of *Blautia*_ASV2217 (with standard error about the mean) in ABIS subjects who later require one of seven chronic diseases (referred to as cases here). The average percent relative abundance of these bacteria is also included. Three cases acquired more than one of these chronic diseases. (SE = standard error about the mean). Here, each disease incidence is considered since four children were diagnosed with more than one disease up to 19 years of age. Rank order abundance of *Blautia*_ASV2217 is shown for each disease followed by the total number of non-zero ASVs in that group of children. By abundance, *Blautia*_ASV2217 ranks in the top 1% to 6·3% of the most abundant taxa in the gut microbiome of cases. NA = not applicable.

| Disease | no. cases | Mean age diagnosis<br>± std dev | age range | % female | % ASV_2217 prevalence | Mean 16S rRNA copies/g (± SE) | % relative abundance | rank order abundance |
| --- | --- | --- | --- | --- | --- | --- | --- | --- |
| any disease | 96 | 12.9 ± 4.7 | 2-19 | 53.7 | 66.7 (64/96) | 428,676 ± 103,974 | 0.91 | 29 (2,461, 1.2%) |
| autism | 31 | 12.6 ± 3.8 | 4-18 | 40.0 | 71.9 (23/32) | 418,618 ± 174,818 | 1.03 | 30 (1,229, 2.4%) |
| all autoimmune | 65 | 12.8 ± 4.9 | 2-18 | 61.5 | 63.1 (41/65) | 426,668 ± 129,851 | 0.86 | 19 (1,945, 1.0%) |
| celiac disease | 29 | 10.0 ± 5.6 | 2-19 | 65.5 | 62.1 (18/29) | 336,431 ± 128,299 | 0.78 | 37 (1,316, 2.8%) |
| hypothyroidism | 6 | 15.3 ± 1.6 | 13-17 | 100.0 | 57.1 (4/7) | 1,169,717 ± 981,935 | 1.05 | 25 (358, 5.9%) |
| Crohn's disease | 6 | 16.0 ± 1.4 | 14-18 | 33.3 | 85.7 (6/7) | 194,455 ± 143,450 | 0.69 | 33 (526, 6.3%) |
| JIA | 11 | 13.2 ± 5.2 | 4-18 | 63.6 | 54.5 (6/11) | 541,732 ± 253,615 | 1.05 | 27 (704, 3.8%) |
| type 1 diabetes | 13 | 13.1 ± 3.2 | 7-17 | 38.5 | 71.4 (10/14) | 707,118 ± 496,857 | 0.78 | 19 (741, 2.6%) |
| Healthy controls | 1,645 | NA | NA |  | 0 | 0 | 0 | NA |

Comparing autoimmune subjects (n = 56) to their matched controls (n = 102), 99 ASVs in 28 genera were identified. In the smaller subset of autism subjects (n = 24) compared to their matched controls (n = 37), only 44 ASVs in 17 genera were identified as significant. Interestingly, combining all disorder developments, both autoimmune and autism, resulted in many overlapping ASVs. 94 ASVs in 28 genera were found to be significantly different comparing subjects that developed a disorder (n = 80) and subjects that remained healthy (n = 139).

### Microbial community differences between cases and controls

Microbial community composition distances were significantly different between cases and matched controls regardless of the distance measure used: binomial, Bray-Curtis, or Jaccard (PERMANOVA: R^2^ = 0.01567, F Model = 1.7197, p-value = 0.001, distance metric: binomial). Healthy matched control microbial communities also differed significantly between the autistic group and the autoimmune group individually (Supplementary Table 1). The microbial communities of autism and autoimmune subjects did not differ significantly from each other.

Heat maps of ASV prevalence between cases and control illustrate ASV exclusivity using controls matched by HLA genotype and location (Figure 1A) and randomly chosen controls (Figure 1B). Given the high number of controls, controls-exclusive ASV prevalence was never higher than 25.9% (Table 2). Cases-exclusive ASV prevalence is as high as 66.7% likely because of the lower number of cases.

**Table 2.** List of all bacterial strains found exclusively in stool of one-year old ABIS subjects who were either cases (acquired one of seven chronic diseases during the first 19 years of life) or controls (remained healthy for 19 years). The ABIS cohort includes 96 subjects with future disease and 1,645 with no future disease up to 19 years of age. All gut microbiome biomarkers (GMBs) differ significantly between the two groups by Chi Square with p-values mostly at or very near zero. To calculate odds ratio, an expected level of prevalence was determined by randomizing the dataset 10 times and creating an artificial set of “cases” and “controls” for each randomization (Supplementary Table 3). The expected prevalence levels for the artificial “cases” varied from 1.8% to 1.98% when case prevalence was zero. The expected prevalence levels for the artificial “controls” varied from 2.84% to 4.40% to 1.98% when control prevalence was zero. Rank order abundance of each non-zero ASV is listed for cases (out of 2,461 ASVs) and controls (out of 14,857 ASVs) along with the % of ASVs that rank higher in cases or controls.

| <u>Bacterium</u> | <u>% prevalence</u> |  | <u>odds ratios</u> |  | <u>Chi-Square</u><br><u>p-value</u> | <u>rank order</u><br><u>abundance</u> |
| --- | --- | --- | --- | --- | --- | --- |
|  | <u>cases</u> | <u>controls</u> | <u>cases</u> | <u>controls</u> |  |  |
| *Blautia*_ASV2217 | 66.7 | 0.0 | 33.69 | 0.0297 | 0 | 29 (1.2%) |
| *Fusicatenibacter*_ASV6284 | 35.8 | 0.0 | 20.22 | 0.0495 | 0 | 172 (7.0%) |
| *Blautia*_ASV2223 | 28.4 | 0.0 | 16.06 | 0.0622 | 0 | 119 (4.8%) |
| *Bifidobacterium*_ASV2142 | 0.0 | 6.7 | 0.423 | 2.36 | 0.0216 | 58 (0.4%) |
| *Bifidobacterium*_ASV1984 | 0.0 | 9.9 | 0.296 | 3.38 | 4.65e-5 | 38 (0.2%) |
| *Faecalibacterium*_ASV5640 | 0.0 | 12.2 | 0.247 | 4.05 | 1.25e-7 | 156 (1.1%) |
| *Faecalibacterium*_ASV5639 | 0.0 | 14.2 | 0.220 | 4.54 | 3.91e-10 | 137 (0.9%) |
| *Akkermansia*_ASV701 | 0.0 | 16.3 | 0.200 | 4.99 | 5.54e-13 | 173 (1.2%) |
| *Collinsella*_ASV4246 | 0.0 | 18.9 | 0.178 | 5.61 | 0 | 82 (0.6%) |
| *Faecalibacterium*_ASV5726 | 0.0 | 21.6 | 0.163 | 6.13 | 0 | 129 (0.9%) |
| *Clostridium*_ASV3940 | 0.0 | 22.6 | 0.168 | 5.96 | 0 | 200 (1.3%) |
| *Collinsella*_ASV4245 | 0.0 | 25.9 | 0.161 | 6.23 | 0 | 52 (0.4%) |
| *Romboutsia*_ASV9859 | 0.0 | 25.9 | 0.170 | 5.87 | 0 | 196 (1.3%) |

**Figure 1.**
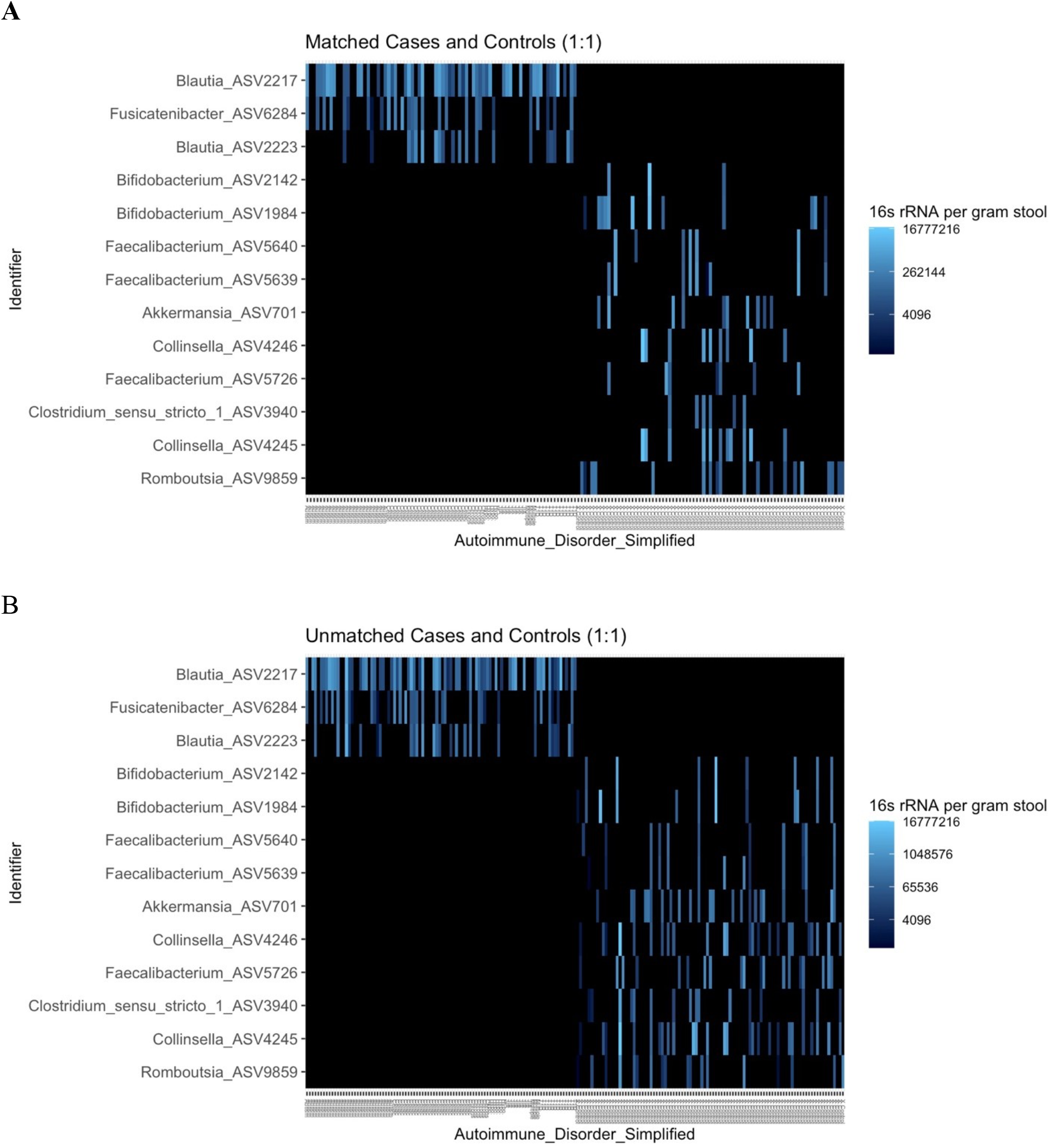
Heat maps illustrating the distribution of 3 ASVs that are exclusive to cases and the 10 ASVs that are found only in controls. **A)** Cases (those with future disease on the left of the x-axis) are matched with one control (no disease to the right) by HLA genotype and geography as much as possible by municipality (cases n= 58), county (cases n = 19) region (cases n = 1), or all of Sweden (cases n = 5); **B)** cases on the left are unmatched with a randomly selected set of controls on the right.

Strikingly, *Blautia_*ASV2217, was observed exclusively in two-thirds of the 96 cases but was completely absent in the 1,645 controls (Tables 1 and 2). Analysis via qPCR of all samples using universal bacteria primers showed that the average abundance of *Blautia*_ASV2217 was over 428,000 copies per gram of stool per sample with a relative abundance of just over 1% and a rank order abundance of 29 (Table 1). Therefore, given all of the bacteria present in stool, *Blautia*_ASV2217 is likely not a trivial player in this ecosystem. Two other ASVs were also found to be exclusive to cases, *Blautia_*ASV2223, and *Fusicatenibacter*_ASV6284 (Table 2).

Ten gut bacterial ASVs were exclusive to controls and were entirely absent in children who later developed autoimmune diseases or autism (Table 2). The controls-exclusive ASVs include two *Bifidobacterium* strains, three *Faecalibacterium* strains, two *Collinsella* strains, and *Akkermansia, Clositridium*, and *Romboutsia* strains. *Romboutsia*_ASV9859 was identified in 25.9% of control cases and is of particular interest due to its juxtaposition to *Blautia*_ASV2217 when considering genetic risk patterns for autoimmunity.

### Effect of HLA host genetics on colonization by cases- and controls-specific ASVs

The presence of *Blautia_*ASV2217 in cases was strongly correlated with (DR4)-DQA1*03-DQB1*03:02 (DR4-DQ8) allele dosage (Figure 2A). However, there was no association between DR4-DQ8 allele dosage and either the abundance or relative abundance of *Blautia*_ASV2217. This suggests that DR4-DQ8 influences initial colonization by *Blautia*_ASV2217, but once colonization occurs, it has no effect on the population size or proportion of this bacterium within the gut. Colonization of *Fusicatenibacter*_ASV6284 was also correlated to the presence of the DR4-DQ8 allele, but abundance after initial colonization is unaffected (Supplemental Table 1, Figure 2B). In other words, DR4-DQ8 may serve as a genetic gatekeeper for *Blautia*_ASV2217 and *Fusicatenibacter*_ASV6284 gut colonization (Figure 2B). The DR4-DQ8 allele was found in cases from all diseases at varying frequencies, but differences were not significant for lack of power. (DR1/10)-DQB1*05:01 (DR1-DQ5) is also correlated with *Fusicatenibacter*_ASV6284 colonization (Figure 2B). All associations listed in Figure 2 have a p-value of 0.05 or less. Precise p-values of all associations are in Supplementary Table 1.

**Figure 2.**
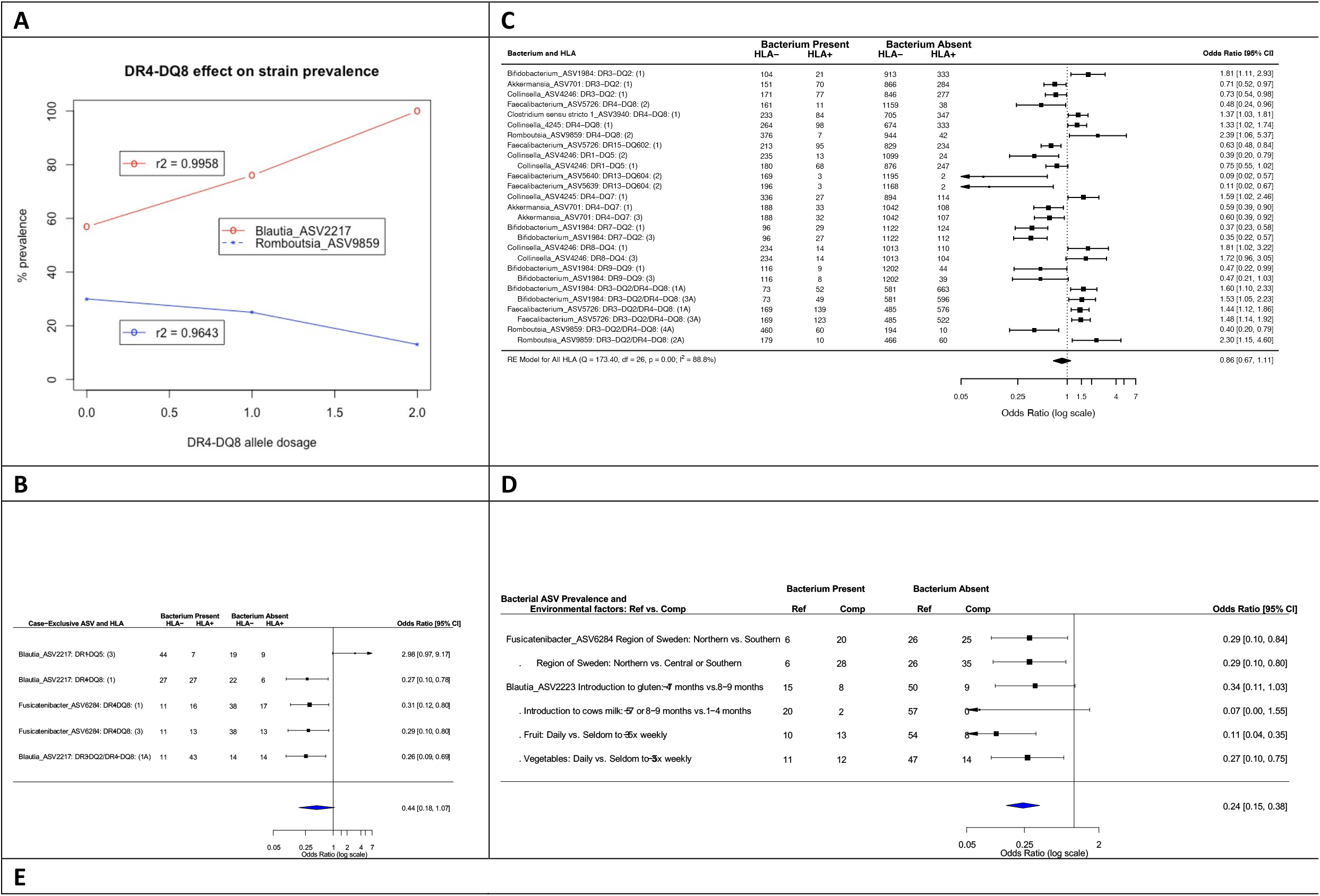
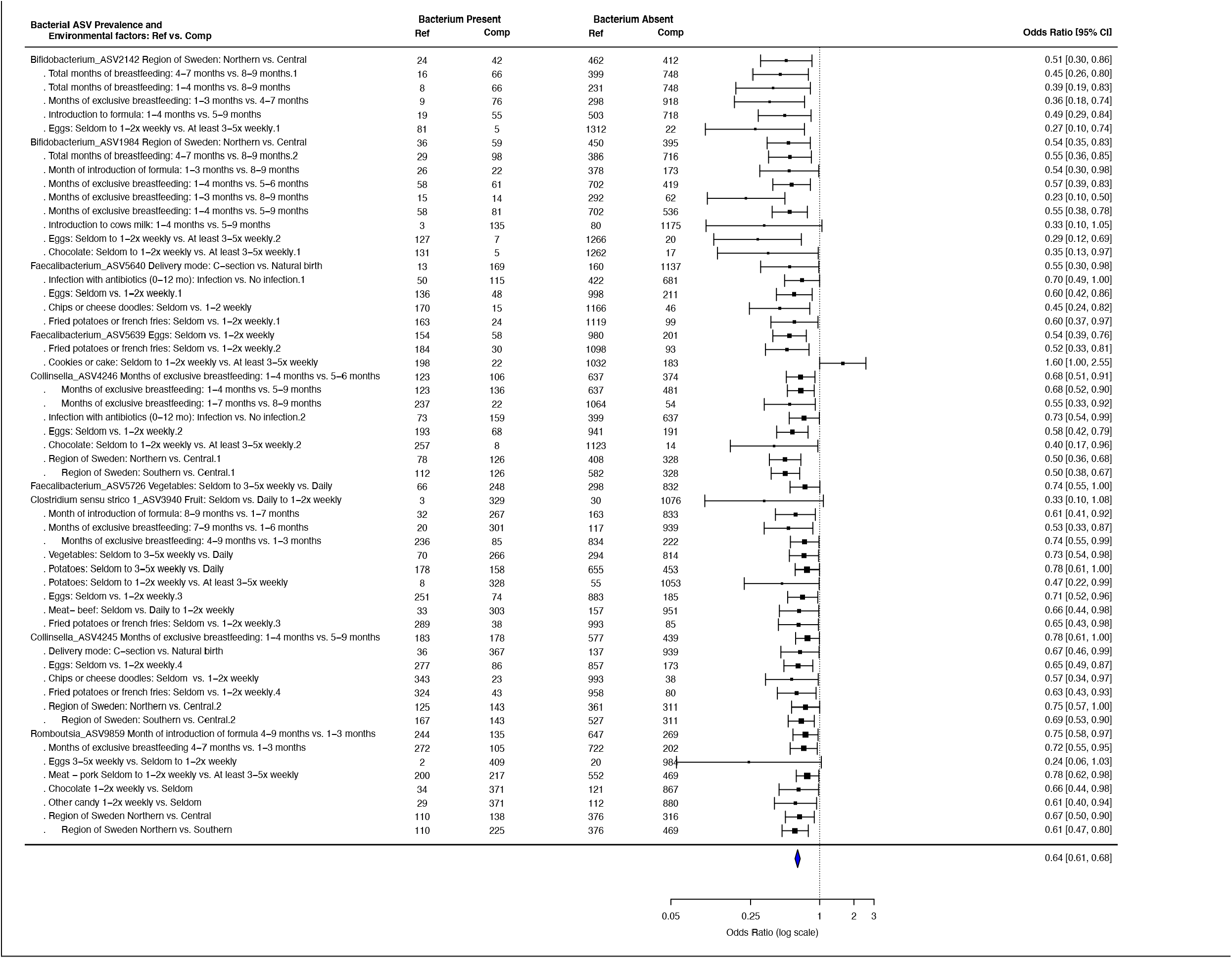
HLA (A,B,C) and environmental factors (D,E) associated with ASVs prevalent exclusively in cases (B,C) or controls (D,E). A) prevalence of case-exclusive *Blautia* ASV_2217 and controls-exclusive *Romboutsia*_ASV9859 are positively (r^2^ = 0.9958) and negatively (r^2^ = 0.9643) correlated DR4-DQ8 allele dosage; *Blautia* ASV_2217 and *Romboutsia*_ASV9859 are the most prevalent ASVs that are exclusive to cases or controls, respectively; **B-E)** Comparisons for the HLA alleles are as follows: (1) compares as HLA-, i.e., those (-/-) with HLA+, i.e., those (-/+ or +/+) for the allele indicated, (2) compares (-/- or -/+) with (+/+) for the allele, (3) compares (-/-) with (-/+) for the allele. The combined HLA models consider DR3-DQ2 and DR4-DQ8 prevalence and are as follows: (1A) compares HLA-, i.e., those homozygous (-/-) at both alleles with those who have at least one of the two alleles (+/- or +/+), (2A), compares those with at least one allele of either DR3-DQ2 or DR4-DQ8 with those who are homozygous (+/+) at both, (3A) compares those who are (-/- for both DR3-DQ2 and DR4-DQ8) with those who are homozygous +/+ for *either* DR3-DQ2 or DR4-DQ8, and (4A) compares those who are homozygous (-/-) for both alleles with those who are homozygous (+/+) at both alleles. For comparisons of ASVs with environmental factors, reference (Ref) and comparison (Comp) are shown for each association, with odds ratios (OR) calculated from the reference group. ORs below or above unity indicate that the bacterium is less or more likely to be present in those categorized as the reference environmental factor. In C), prevalence of case-exclusive *Blautia* ASV_2217 and controls-exclusive *Romboutsia*_ASV9859 are positively (r^2^ = 0.9958) and negatively (r^2^ = 0.9643) correlated DR4-DQ8 allele dosage; *Blautia* ASV_2217 and *Romboutsia*_ASV9859 are the most prevalent ASVs that are exclusive to cases or controls, respectively. All associations listed here are significant with a p-value less than 0.05. Precise p-values for each association are listed in Supplementary Table 1.

The opposite HLA effect was seen with controls-exclusive ASVs (Figure 2A,B). Many of these ASVs are members of bacterial genera that are typically beneficial to gut health such as *Bifidobacterium* and *Faecalibacterium* (Table 2).^22,23^ Gut colonization of these bacteria appears discouraged by autoimmunity-associated high-risk haplotypes such as DR3-DQ2 ((DR3)-DQA1*05-DQB1*02) or DR4-DQ8 but encouraged by other alleles such as DR7-DQ2 ((DR7)-DQA1*02:01-DQAB1*02) and DR9-DQ9 ((DR9)-DQA1*03-DQB1*03:03) (Fig. 2C, Supplemental Table 1). Hence, certain HLA alleles may limit colonization by beneficial bacteria, suggesting that the efficacy of probiotic restoration of a healthy microbiome may be very difficult in subjects at high risk for autoimmune diseases or autism. An understanding of how HLA proteins interact with certain bacteria may be improve the benefit of probiotics. Interestingly, a counteracting effect was found where DR4-DQ8 encouraged colonization by *Blautia*_ASV2217, the most prevalent cases-exclusive ASV, but discouraged colonization by *Romboutsia*_ASV9859, the most prevalent controls-exclusive ASV (Figure 2A-C).

### Effect of non-genetic factors on colonization by cases- and controls-specific ASVs

In addition to host genetics, environmental factors that may influence gut microbiome composition were considered for effect on gut colonization by the ASVs of interest. These factors include approximate location of the residence, diet, antibiotic use during pregnancy, mode of delivery, breast feeding, infections, gender, siblings, and pets. No effect of sex of the child, number of siblings, or pets was observed. However, strong associations with diet and maternal factors were identified (Figure 2D,E, Supplementary Table 1).

Several dietary factors were significantly associated with the colonization of *Blautia*_ASV2223, a case-exclusive ASV not controlled by HLA genetics (Figure 2). Presence of *Blautia*_ASV2223 was associated with introduction of gluten later in infancy (8-9 months vs. 4-7 months), *χ2*(1)=3.8, *p*=0·05, and reduced fruit and vegetable consumption. Conversely, presence of most of the controls-exclusive ASVs was associated with increased frequency of egg consumption by the infant (Figure 2D, Supplementary Table 1). Where control-specific colonization was influenced by mode of delivery, it was always on the side of natural childbirth rather than C-section (Figure 2). Similarly, longer periods of exclusive breast feeding were associated with colonization of two control-specific ASVs (Figure 2D).

### Characteristics of cases-exclusvive *Blautia*_ASV2217 and *Fusicatenibacter*_ASV6284

Fascinatingly, gut bacterial strain, *Blautia*_ASV2217, was present at one year of age in two-thirds of children who subsequently acquired chronic disease up to 18 years later but is completely absent in the 1,645 children who remained healthy during that time frame. Also, the presence or absence of this bacterium is largely driven by HLA allele DR4-DQ8—molecules that confers high genetic risk for type 1 diabetes, celiac disease, and other autoimmune disorders.^24^ HLA-associated *Blautia_*ASV2217 was also associated with autism, which has been associated with certain HLA alleles but represents a disorder that is not considered a classical autoimmune disease.

The partial 16S rRNA sequence of *Blautia*_ASV2217 was 99.8% identical to or the equivalent 400-base region of the 16S rRNA gene from *Blautia wexlerae* H020 (Supplementary Table 2). The biology of *B. wexlerae* is understudied. This species was first identified from stool samples recovered from children in a late-onset autism study.^25,26^ After its initial discovery, several studies have shown that *B. wexlerae* is enriched in non-obese subjects with insulin resistance.^27,28^ *Blautia wexlerae* was positively correlated with fasting glucose and insulin, HOMA-IR, HOMA-β and triglyceride concentrations,^29^ is a starch degrader,^30^ and has been associated with increased consumption of whole grains.^30^ In a study of the gut microbiota of obese and non-obese adults, *B. wexlerae* was one of five bacterial species shown to be associated with nonobese subjects.^28^ Another study confirmed a decreasing incidence of *B. wexlerae* in the gut microbiota of obese children, especially those also suffering insulin resistance.^27^ This study also demonstrated that *B. wexlerae* exerted anti-inflammatory effects on peripheral blood mononuclear cells in vitro. *Blautia wexlerae* was enriched in metabolically healthy subjects when compared to prediabetes and insulin-resistant adults.^28^ Previous reports associating *B. wexlerae* with anti-inflammatory responses cannot exclude the possibility that other strains of the same species, such as *Blautia*_ASV2217, may have very different effects. In addition, the stool samples from the ABIS cohort contained many ASVs related to *wexlerae*; of those, only *Blautia*_ASV2217 was found exclusively in cases.

The second most prevalent case-specific bacterium was *Fusicatenibacter*_ASV6284 that has 99.8% identity to the 16S rRNA gene of *Fusicatenibacter saccharivorans* V030. *Fusicatenibacter saccharivorans* was first isolated in healthy adults from human feces^32^ and ferments glucose to lactic acid, formic acid, acetic acid, and succinic acid.^33^ In subjects with ulcerative colitis, the relative abundance of *F. saccharivorans* decreased in the gut compared to quiescent or healthy controls.^33^ Heat-killed *F. saccharivorans* induced IL-10 production by lamina propria mononuclear cells from both colitis model mice and human colitis subjects.^33^ Cirrhosis was associated with significantly lower abundance of *F. saccharivorans*, which became more profound as the disease progressed and was found to have consistent association with short chain fatty acid production.^34^ *Fusicatenibacter saccharivorans* had significantly lower abundance in rheumatoid arthritis in comparison to osteoarthritis patients.^35^ Thus, *F. saccharivorans* is typically more common in healthy subjects than in those with disease. This is the first work to associate a specific strain of this species to future autoimmunity or autism.

### Characteristics of controls-exclusive ASVs from the genera *Bifidobacterium, Faecalibacterium, Collinsella, Akkermansia*, and *Romboutsia*

Five of the 10 strains found exclusively in controls are likely members of two species well known to contribute to gut health: *Bifidobacterium longum* subsp. infantis or *Faecalibacterium prausnitzii*.^22,23^ Two different strains of *Collinsella aerofaciens*, one with 100% and the other with 99·8% identity to strain JCM10188, make up 20% of the controls-exclusive ASVs. Notably, *F. prausnitzii* and *C. aerofaciens* are butyrate producers.^36,37^ Butyrate is well known as an anti-inflammatory that contributes to gut epithelial layer integrity.^38^ This is seen as confirmatory evidence that our decision to take a prevalence approach with these data is consistent with past findings on gut health.^22,23,34,37^ These five strains may be contributing to the gut health of ABIS children there by reducing the likelihood of future disease. *Akkermansia*_ASV701 found in 16.3% of control subjects was determined to be *Akkermansia municiphila*. Several studies in humans and in mice suggest that *A. municiphila* strains confer anti-inflammatory effects.^39^ As inflammation is associated with the diseases of interest in this work,^40,41^ the association of *A. municiphila* with controls fits the underlying biology of these diseases.

Strains in the *Romboutsia* genus were associated with low genetic risk for type 1 diabetes in our previous work.^17^ Here, we show that a strain of *Romboutsia* is significantly associated with those controls who never acquire disease. The partial 16S rRNA sequence of *Romboutsia*_ASV9859 was 99.5% identical to *Romboutsia timonensis* strain 1AT-D10-92 (Supplemental Table 2). This strain is also more prevalent in controls than any other control-exclusive bacterium in the dataset. These results suggest that *Romboutsia* strains may be important as probiotics to correct microbiome dysbiosis and protect against future disease. Little is known regarding the biology of *Romboutsia*, but recent work in murine models showed that vitamin C or sodium butyrate administration can increase levels of *Romboutsia* in the gut.^42,43^ Previous work has also shown that increased metabolic potential for butyrate production in the gut is associated with autoimmunity for type 1 diabetes.^5,44^ Thus, while speculative, *Romboutsia* might play an important role in gut integrity, preventing antigens from passing through the gut epithelial layer.

### Contrasting DR4-DQ8 influence on *Blautia*_ASV2217 and *Romboutsia*_ASV9859

The presence or absence of Blautia_ASV2217 was observed to be largely driven by HLA-DR4-DQ8 (Figure 2A). Allele dosage of DR4-DQ8 has the opposite effect on control-exclusive *Romboutsia*_ASV9859, as its prevalence is negatively correlated with DR4-DQ8 (Figure 2A,C). *Romboutsia*_ASV9859 was able to colonize exclusively one-fourth of the 1645 controls, and never in any of the 96 cases (Table 2). Results presented here suggest that DR4-DQ8 can serve as a gatekeeper for the two most prominent biomarkers described here. Specifically, when homozygous, this allele limits the presence of *Romboutsia*_ASV9859, a member of the genus previously associated with high genetic risk for type 1 diabetes,^17^ and encourages colonization of *Blautia*_ASV2217.

## Conclusions

These results have three significant implications: 1) certain class II HLA alleles may serve as bacterial-community gatekeepers early in life, altering microbiome composition and potential for dysbiosis and inflammation; 2) dysbiosis and inflammation in the gut during infancy may be a common precedent to all five autoimmune diseases and autism given certain environmental triggers; and 3) HLA gatekeeping of certain bacteria may prevent gut colonization of beneficial strains in those at-risk individuals who could most benefit for probiotic therapy.

The DR4-DQ8 allele is strongly associated with autoimmune diseases and is the highest risk associated allele for type 1 diabetes.^45^ DQ8 is known to bind many peptides^24^ and, as a result, induce Th17 cell presentation leading to secretion of the IL-17 and IL-23 cytokines.^47^ In turn, these cytokines assist in clearing infections, particularly viral infections.^46^ Results presented here suggest that DR4-DQ8 can serve as a gatekeeper for the presence or absence of bacteria in the gut microbiome.

The evidence presented here suggests gut microbiome dysbiosis is established at one year of age and that this dysbiosis is associated with autoimmune disease or autism up to 18 years later. Dysbiosis appears to be similar across all six diseases. Some of this dysbiosis is controlled by HLA genetics, particularly DR4-DQ8 and DR3-DQ2, but much of it may be regulated by maternal or infant diet. Dysbiosis can lead to inflammation and gut leakiness leading to the diseases examined here. Other factors then trigger the specific disease acquired by the child. Finding approaches to identify and correct this dysbiosis and monitor it across time may prevent future disease. The biomarkers identified here may help in an approach to rapidly identify dysbiosis.

High risk HLA alleles for autoimmune disorders may be preventing beneficial bacteria from inhabiting the gut. Indeed, six of the 10 controls-exclusive ASVs are members of bacterial species have been associated with gut health including *Akkermansia* muciniphila,^48^ *Bifidobacterium longum* subsp. infantis,^49^ and *Faecalibacterium prausnitzii*^50^ (Supplementary Table 1).

If that is the case, probiotics may not adjust this dysbiosis for failure to colonize the gut. Hence, understanding the mechanisms underlying HLA interactions with gut bacteria is crucial to long-term clinical solutions. Long-term diet modifications should also be considered to reduce a large portion of gut dysbiosis, such as more frequent egg consumption.

## Methods

### Sample Collection

This study is in partnership with the ABIS cohort (All Babies in Southeast Sweden). This was a prospective population-based cohort that invited all mothers of children born from October 1, 1997 to October 1, 1999. Parents gave their informed consent after either oral, written, or video information was provided. Participating mothers completed questionnaires and diaries for the first year of life, which included but were not limited to information on antibiotic use, age of dietary introductions, duration of breastfeeding, environmental factors, and more. Stool samples were collected from the diapers of infants using provided sterile spatula and tubes. These samples were labeled with unique identifiers, and immediately frozen after collection. Samples collected at home were transported frozen, to the WellBaby Clinic. Samples were shipped to the University of Florida in a frozen state and were stored at -80°C until extraction.^17,21^

### Institutional Review Board Approval

The ABIS-study has ethical approvals from the Research Ethics Committees of the Faculty of Health Science at Linköping University, Sweden, Ref. 1997/96287 and 2003/03-092 and the Medical Faculty of Lund University, Sweden (Dnr 99227, Dnr 99321) as described previously.^17^ The microbiome analysis performed at the University of Florida was approved by the University of Florida’s Institutional Review Board as an exempt study IRB201800903.

### Diagnoses

Diagnoses of ABIS children were obtained from the Swedish National Diagnosis Registry. Juvenile idiopathic arthritis (JIA) diagnoses are controlled in casebooks as part of the PhD work of one of the authors (EK) and are very strict and reliable. Type 1 diabetes, Crohn’s disease, and hypothyroidism were diagnosed by standard procedure by child’s pediatrician. Prior to the age of 10, celiac disease was diagnosed based on transglutaminase antibodies and confirmed by gut biopsies. Over age 10, biopsies were done when transglutaminase antibodies were less than 10 times above the healthy range. Autism diagnoses have been confirmed by a specialist in child psychiatry.

### HLA Genotyping

HLA-DR/DQ genotypes associated with risk and protection were defined using typing for HLA-DQB1 and informative -DQA1 and DRB1 alleles for deducing the presence of common European HLA-DR-DQ alleles variously associated with disease risk using sequence specific hybridization with lanthanide labeled oligonucleotide probes.^51^ For the chi square analyses, both presence of the alleles and heterozygosity were considered. Distribution of HLA alleles throughout the cohort are listed in Supplementary Table 5.

### Microbiome analysis

DNA extraction, 16S rRNA barcoded PCR, and V3-V4 16S rRNA Illumina sequencing were performed as described previously.^17^ Demultiplexed sequences processed into amplicon sequencing variants (ASVs) using the DADA2 package in R.^52^ Reads were truncated to a length of 425 nucleotides, after seeing the read quality drop sharply through the “plotQualityProfile” graph. Sequences filtered to allow for no ambiguous nucleotides, a maximum expected error rate of 1, truncation after encountering a base with Q-score 11, and the removal of PhiX reads. Chimeras were removed using the “removeBimeraDenovo” function using the default “consensus” method. Taxonomy was assigned using the SILVA 138 database.^53^ Of the 15,519 ASVs represented in the cohort (N=1,741), 6,057 had counts ≥ 2. The average counts per sample were 32,613 with a maximum of 446,583 counts.

ASVs were first labeled by sorting the ASVs based on known genus and appending unique sequential numbers to the genus. Differentially abundant ASV between disorder development groups were determined using DESeq2 using the Wald Significance testing and “local” dispersion fitting. Only sequences with padv < 0.01 were further visualized using the plot_heatmap function from the phyloseq package in R.^54^ ASVs identified as being significant were then run through NCBI’s blast, to determine any known information about the sequence.

Differential abundances between cases and controls at the species, genus, and family level were obtained for the filtered ASVs using edgeR with an FDR correction.^55^ Controls missing geographical data were excluded. As 83% of these 277 filtered reads have not yet been assigned to the species level in the SILVA database, the original sequences for any ASVs within those significant *undefined* species of a particular genus were then referenced against the NCBI BLAST database to determine homology.

### Matching Cases to Healthy Controls

At one year of age, 1,729 out of 1,741 subjects had a known municipality. These municipalities were then grouped into either established counties of Sweden, or regions based on general location. A map of the area was constructed previously using the R-packages Swemaps and ggplot2.^17,56^ The longitude and latitude of the municipalities were found online and used as place markers for the municipalities.

Of the 1,741 subjects, 96 had Autoimmune or Autism, 82 of which also had a listed HLA Genotype and Municipality. These 82 subjects were matched to healthy controls, those without know Autoimmune (n = 56) or Autism (n = 24) development, based on Genotype and nearest geography. Controls were first matched on Municipality. If there were no matches, the search was expanded to county, region, and all of Sweden. Eighty subjects (two had unique genotypes and were unable to be matched DR14-DQ503/DR15-DQ602 and DR5-DQ7/DR14-DQ5) were matched with 139 controls.

## Statistical analysis

Alpha diversity was determined through the plot richness function of the phyloseq package as well as the ggplot2 package in R.^55^ Statistical testing of the group means was performed using the stat_compare_means function, from the ggpubr package, using both “kruskal.test” and “wilcox.test”. Alpha diversity by region and case/control was calculated using the rarefied, unfiltered reads in MicrobiomeAnalyst.^57^ The 1,741 samples were rarefied to an equal sequencing depth based on the minimum library size (3,507). For these calculations, samples missing geography were excluded. To compare diversity measures across cases and controls, controls were further classified by region (Central, Northern, Southern Sweden) and Kruskal-Wallis tests were conducted on the alpha diversity measures across the four groups.

Microbial community composition distances (binomial, Bray-Curtis, Jaccard) were compared between cases and matched controls with PERMANOVA from the adonis R package. Differences in microbiome composition by the development of Autoimmune Diseases or Autism were tested for using the permutational multivariate analysis of variance (PERMANOVA) test through the “adonis” function in the vegan R package, and the “pairwise.adonis2” function in the PairwiseAdonis R package. All PERMANOVA tests were performed with the default 999 permutations. P-value results from statistical testing were corrected for false discovery rate (FDR) using the Benjamin–Hochberg method. A low-count filter removed 5765 ASVs for which at least 10% of the counts did not equal or exceed 4. A low-variance filter removed 15 additional ASVs with low variance as measured by the inter-quartile range (ICR) and a 5% cutoff. The filtered counts were centered-log ratio transformed in MicrobiomeAnalyst and were not rarefied for this approach to avoid significant data loss.

Chi square tests were run in IBM SPSS on the case-exclusive or control-exclusive ASVs identified by DESeq2^58^ (ASV level) or edgeR (species level) within the respective cohorts to determine whether the presence or absence of the bacterium was associated with genetic risk for autoimmunity, lifestyle, or environment, and an odds ratio calculated with 95% confidence intervals using the metaphor package in R. Samples with missing data were not considered in the calculations.

## Data Availability

16S RNA data will soon be submitted to the Sequence Read Archive at NCBI.

## Acknowledgements

We thank Ingela Johansson, Gosia Smolinska and Åshild Faresjö from Linköping University for their help with sample collection and distribution as well as with providing the authors with register data.

