## Supplemental Table 1 for "Adolescent autism and autoimmune diagnoses linked to infant gut bacteria whose prevalence is associated with at-risk genetics and/or diet": Supplementary_Table_1.docx

**Supplementary Table 1. HLA alleles, dietary, and geographic associations with the three case-specific and 10 control-specific ASVs.** Results of chi-square test of independence for presence/absence of the bacterium in either cases or controls. Where df > 1, the pairwise p value is indicated next to the factor.

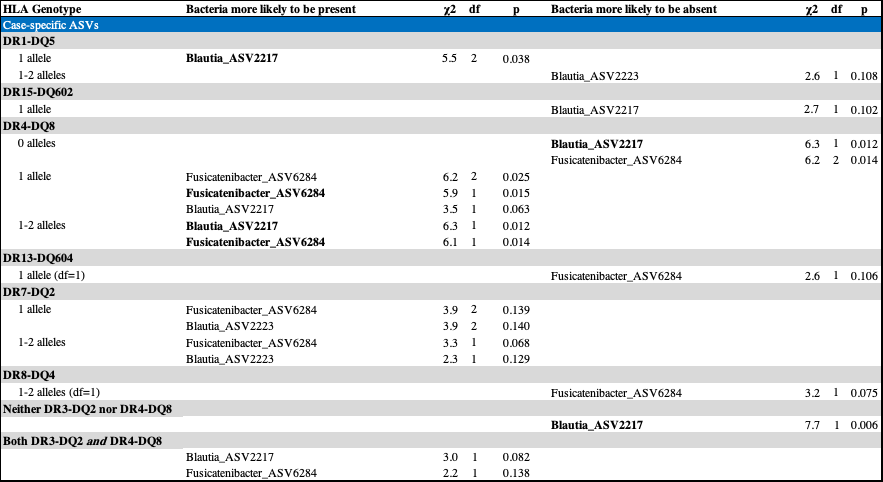

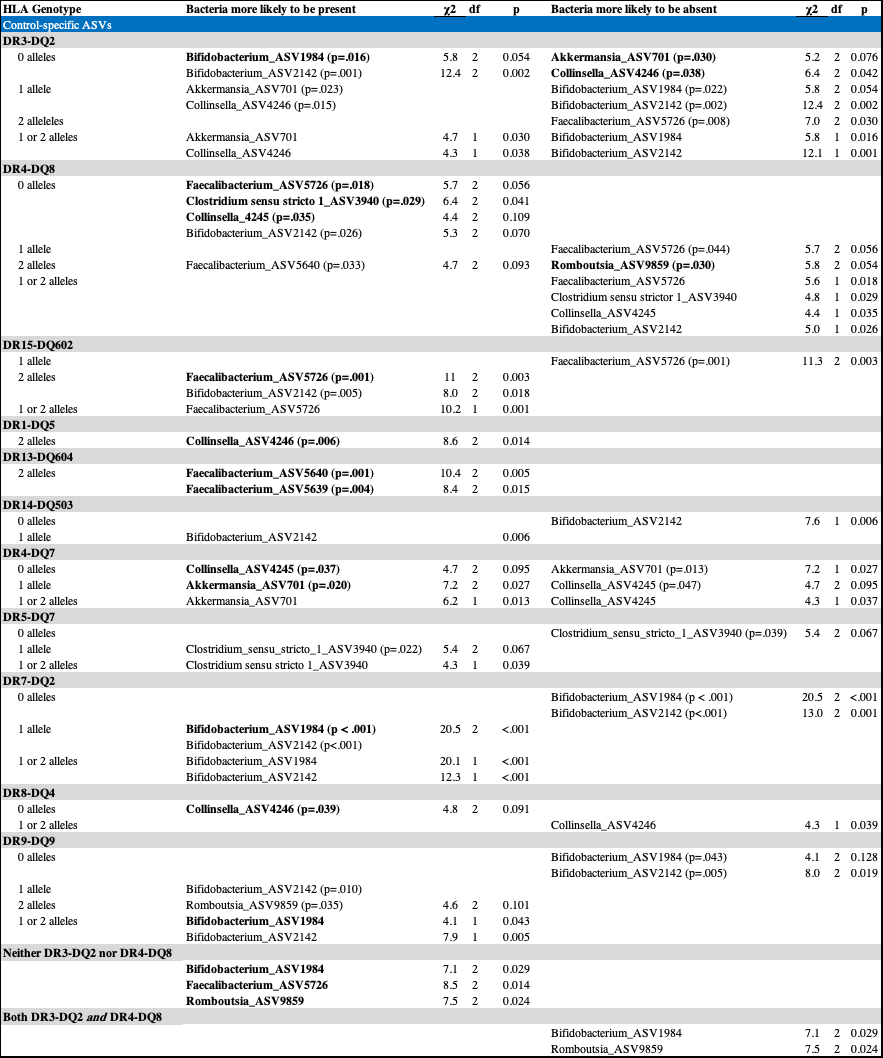

**Supplementary Table 2. HLA alleles, dietary, and geographic associations the three case-specific and 10 control-specific ASVs.** Results of chi-square test of independence for presence/absence of the bacterium in either cases or controls. Where df > 1, the pairwise p value is indicated next to the factor.

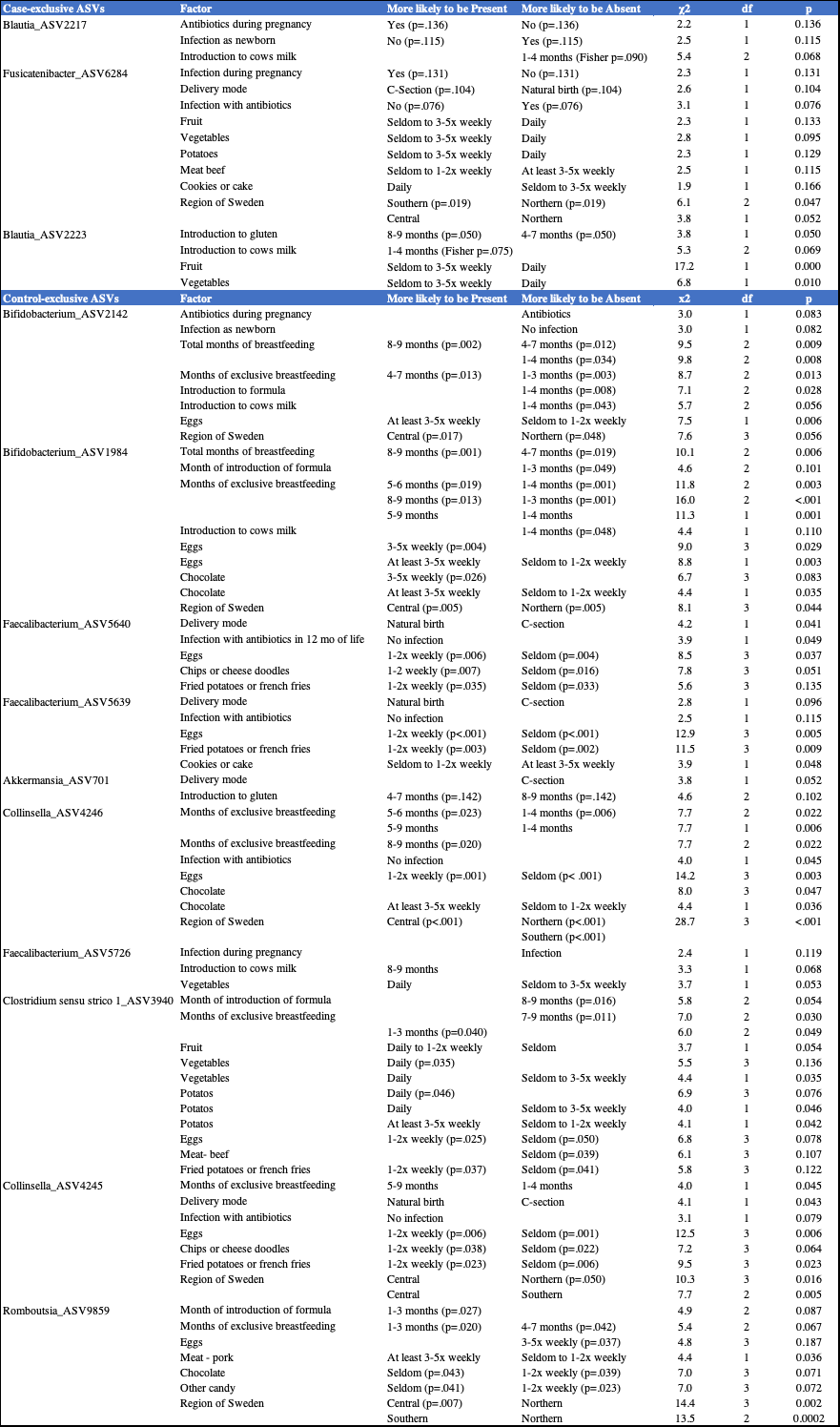
