## Supplemental Table 2 for "Adolescent autism and autoimmune diagnoses linked to infant gut bacteria whose prevalence is associated with at-risk genetics and/or diet": Supplementary_Table_2.pdf

Supplementary Table 1. Closest cultured relatives of all bacterial strains found exclusively in stool of one-year old ABIS subjects who were either cases (acquired one of seven chronic diseases during the first 19 years of life) or controls (remained healthy for 19 years).

| Bacterium | closest cultured relative | % identity |
|---|---|---|
| *Blautia*_ASV2217 | *Blautia wexlerae* HC20 | 99.8 |
| *Fusicatenibacter*_ASV6284 | *Fusicatenibacter saccharivorans V030* | 99.8 |
| *Blautia*_ASV2223 | *Blautia massiliensis* CSUR P2132 | 99.8 |
| *Bifidobacterium*_ASV2142 | *Bifidobacterium longum* subsp. infantis strain Bi-26 | 99.1 |
| *Bifidobacterium*_ASV1984 | *Bifidobacterium longum* subsp. infantis strain Bi-26 | 99.1 |
| *Faecalibacterium*_ASV5640 | *Faecalibacterium prausnitzii* A2-165 strain JCM 31915 | 99.5 |
| *Faecalibacterium*_ASV5639 | *Faecalibacterium prausnitzii* A2-165 strain JCM 31915 | 99.8 |
| *Akkermansia*_ASV701 | *Akkermansia muciniphila* TL23 gene | 100.0 |
| *Collinsella*_ASV4246 | *Collinsella aerofaciens* strain JCM 10188 | 100.0 |
| *Faecalibacterium*_ASV5726 | *Faecalibacterium prausnitzii* isolate MGYG-HGUT-02545 | 99.5 |
| *Clostridium*_ASV3940 | *Clostridium celatum* G085 | 99.8 |
| *Collinsella*_ASV4245 | *Collinsella aerofaciens* strain JCM 10188 | 99.8 |
| *Romboustia*_ASV9859 | *Romboutsia timonensis* strain 1AT-D10-92 | 99.5 |
