## Supplemental Table 3 for "Adolescent autism and autoimmune diagnoses linked to infant gut bacteria whose prevalence is associated with at-risk genetics and/or diet": Supplementary_Table_3.pdf

Blautia_ASV2217
CAGTGGGGAATATTGCACAATGGGGGAAACCCTGATGCAGCGACGCCGCGTGAAGGAAGAAGTATC
TCGGTATGTAAACTTCTATCAGCAGGGAAGATAGTGACGGTACCTGACTAAGAAGCCCCGGCTAACT
ACGTGCCAGCAGCCGCGGTAATACGTAGGGGGCAAGCGTTATCCGGATTTACTGGGTGTAAAGGGA
GCGTAGACGGTGTGGCAAGTCTGATGTGAAAGGCATGGGCTCAACCTGTGGACTGCATTGGAAACT
GTCATACTTGAGTGCCGGAGGGGTAAGCGGAATTCCTAGTGTAGCGGTGAAATGCGTAGATATTAGG
AGGAACACCAGTGGCGAAGGCGGCTTACTGGACGGTAACTGACGTTGAGGCTCGAAAGCGTGGGGA
GCAAACAGGATTAGATACCCGGGTAGT

Fusicatenibacter_ASV6284
CAGTGGGGAATATTGCACAATGGGGGAAACCCTGATGCAGCGACGCCGCGTGAGCGAAGAAGTATT
TCGGTATGTAAAGCTCTATCAGCAGGGAAGATAATGACGGTACCTGACTAAGAAGCCCCGGCTAACT
ACGTGCCAGCAGCCGCGGTAATACGTAGGGGGCAAGCGTTATCCGGATTTACTGGGTGTAAAGGGA
GCGTAGACGGCAAGGCAAGTCTGATGTGAAAACCCAGGGCTTAACCCTGGGACTGCATTGGAAACTG
TCTGGCTCGAGTGCCGGAGAGGTAAGCGGAATTCCTAGTGTAGCGGTGAAATGCGTAGATATTAGG
AAGAACACCAGTGGCGAAGGCGGCTTACTGGACGGTAACTGACGTTGAGGCTCGAAAGCGTGGGGA
GCAAACAGGATTAGATACCCGGGTAGT

Blautia_ASV2223
CAGTGGGGAATATTGCACAATGGGGGAAACCCTGATGCAGCGACGCCGCGTGAAGGAAGAAGTATC
TCGGTATGTAAACTTCTATCAGCAGGGAAGAAAATGACGGTACCTGACTAAGAAGCCCCGGCTAACT
ACGTGCCAGCAGCCGCGGTAATACGTAGGGGGCAAGCGTTATCCGGATTTACTGGGTGTAAAGGGA
GCGTAGACGGATGGACAAGTCTGATGTGAAAGGCTGGGGCTCAACCCCGGGACTGCATTGGAAACT
GCCCGTCTTGAGTGCCGGAGAGGTAAGCGGAATTCCTAGTGTAGCGGTGAAATGCGTAGATATTAGG
AGGAACACCAGTGGCGAAGGCGGCTTACTGGACGGTAACTGACGTTGAGGCTCGAAAGCGTGGGGA
GCAAACAGGATTAGATACCCGGGTAGT

Bifidobacterium_ASV2142
CAGTGGGGAATATTGCACAATGGGCGCAAGCCTGATGCAGCGACGCCGCGTGAGGGATGGAGGCCT
TCGGGTTGTAAACCTCTTTTATCGGGGAGCAAGCGTGAGTGAGTTTACCCGTTGAATAAGCACCGGCT
AACTACGTGCCAGCAGCCGCGGTAATACGTAGGGTGCAAGCGTTATCCGGAATTATTGGGCGTAAAG
GGCTCGTAGGCGGTTCGTCGCGTCCGGTGTGAAAGTCCATCGCTTAACGGTGGATCCGCGCCGGGTA
CGGGCGGGCTTGAGTGCGGTAGGGGAGACTGGAATTCCCGGTGTAACGGTGGAATGTGTAGATATC
GGGAAGAACACCAATGGCGAAGGCAGGTCTCTGGGCCGTTACTGACGCTGAGGAGCGAAAGCGTGG
GGAGCGAACAGGATTAGATACCCGT

Bifidobacterium_ASV1984
CAGTGGGGAATATTGCACAATGGGCGCAAGCCTGATGCAGCGACGCCGCGTGAGGGATGGAGGCCT
TCGGGTTGTAAACCTCTTTTATCGGGGAGCAAGCGTGAGTGAGTTTACCCGTTGAATAAGCACCGGCT
AACTACGTGCCAGCAGCCGCGGTAATACGTAGGGTGCAAGCGTTATCCGGAATTATTGGGCGTAAAG
GGCTCGTAGGCGGTTCGTCGCGTCCGGTGTGAAAGTCCATCGCTTAACGGTGGATCCGCGCCGGGTA

CGGGCGGGCTTGAGTGCGGTAGGGGAGACTGGAATTCCCGGTGTAACGGTGGAATGTGTAGATATC
GGGAAGAACACCAATGGCGAAGGCAGGTCTCTGGGCCGTTACTGACGCTGAGGAGCGAAAGCGTGG
GGAGCGAACAGGATTAGATACCCGG

Faecalibacterium_ASV5640
CAGTGGGGAATATTGCACAATGGGGGAAACCCTGATGCAGCGACGCCGCGTGGAGGAAGAAGGTCT
TCGGATTGTAAACTCCTGTTGTTGAGGAAGATAATGACGGTACTCAACAAGGAAGTGACGGCTAACT
ACGTGCCAGCAGCCGCGGTAAAACGTAGGTCACAAGCGTTGTCCGGAATTACTGGGTGTAAAGGGA
GCGCAGGCGGGAAGACAAGTTGGAAGTGAAATCTATGGGCTCAACCCATAAACTGCTTTCAAAACTG
TTTTTCTTGAGTAGTGCAGAGGTAGGCGGAATTCCCGGTGTAGCGGTGGAATGCGTAGATATCGGGA
GGAACACCAGTGGCGAAGGCGGCCTACTGGGCACCAACTGACGCTGAGGCTCGAAAGTGTGGGTAG
CAAACAGGATTAGATACCCCTGTAGT

Faecalibacterium_ASV5639
CAGTGGGGAATATTGCACAATGGGGGAAACCCTGATGCAGCGACGCCGCGTGGAGGAAGAAGGTCT
TCGGATTGTAAACTCCTGTTGTTGAGGAAGATAATGACGGTACTCAACAAGGAAGTGACGGCTAACT
ACGTGCCAGCAGCCGCGGTAAAACGTAGGTCACAAGCGTTGTCCGGAATTACTGGGTGTAAAGGGA
GCGCAGGCGGGAAGACAAGTTGGAAGTGAAATCTATGGGCTCAACCCATAAACTGCTTTCAAAACTG
TTTTTCTTGAGTAGTGCAGAGGTAGGCGGAATTCCCGGTGTAGCGGTGGAATGCGTAGATATCGGGA
GGAACACCAGTGGCGAAGGCGGCCTACTGGGCACCAACTGACGCTGAGGCTCGAAAGTGTGGGTAG
CAAACAGGATTAGATACCCCGGTAGT

Akkermansia_ASV701
CAGTCGAGAATCATTCACAATGGGGGAAACCCTGATGGTGCGACGCCGCGTGGGGGAATGAAGGTC
TTCGGATTGTAAACCCCTGTCATGTGGGAGCAAATTAAAAAGATAGTACCACAAGAGGAAGAGACGG
CTAACTCTGTGCCAGCAGCCGCGGTAATACAGAGGTCTCAAGCGTTGTTCGGAATCACTGGGCGTAA
AGCGTGCGTAGGCTGTTTCGTAAGTCGTGTGTGAAAGGCGCGGGCTCAACCCGCGGACGGCACATG
ATACTGCGAGACTAGAGTAATGGAGGGGGAACCGGAATTCTCGGTGTAGCAGTGAAATGCGTAGAT
ATCGAGAGGAACACTCGTGGCGAAGGCGGGTTCCTGGACATTAACTGACGCTGAGGCACGAAGGCC
AGGGGAGCGAAAGGGATTAGATACCCG

Collinsella_ASV4246
CAGTGGGGAATCTTGCGCAATGGGGGGAACCCTGACGCAGCGACGCCGCGTGCGGGACGGAGGCC
TTCGGGTCGTAAACCGCTTTCAGCAGGGAAGAGTCAAGACTGTACCTGCAGAAGAAGCCCCGGCTAA
CTACGTGCCAGCAGCCGCGGTAATACGTAGGGGGCGAGCGTTATCCGGATTCATTGGGCGTAAAGC
GCGCGTAGGCGGCCCGGCAGGCCGGGGGTCGAAGCGGGGGGCTCAACCCCCCGAAGCCCCCGGAA
CCTCCGCGGCTTGGGTCCGGTAGGGGAGGGTGGAACACCCGGTGTAGCGGTGGAATGCGCAGATAT
CGGGTGGAACACCGGTGGCGAAGGCGGCCCTCTGGGCCGAGACCGACGCTGAGGCGCGAAAGCTG
GGGGAGCGAACAGGATTAGATACCCGTGTAG

Faecalibacterium_ASV5726
CAGTGGGGAATATTGCACAATGGGGGAAACCCTGATGCAGCGACGCCGCGTGGAGGAAGAAGGTCT
TCGGATTGTAAACTCCTGTTGTTGAGGAAGATAATGACGGTACTCAACAAGGAAGTGACGGCTAACT
ACGTGCCAGCAGCCGCGGTAAAACGTAGGTCACAAGCGTTGTCCGGAATTACTGGGTGTAAAGGGA
GCGCAGGCGGGAGAACAAGTTGGAAGTGAAATCCATGGGCTCAACCCATGAACTGCTTTCAAAACTG
TTTTTCTTGAGTAGTGCAGAGGTAGGCGGAATTCCCGGTGTAGCGGTGGAATGCGTAGATATCGGGA
GGAACACCAGTGGCGAAGGCGGCCTACTGGGCACCAACTGACGCTGAGGCTCGAAAGTGTGGGTAG
CAAACAGGATTAGATACCCGTGTAGT

Clostridium_ASV3940
CAGTGGGGAATATTGCACAATGGGGGAAACCCTGATGCAGCAACGCCGCGTGAGTGATGACGGCCT
TCGGGTTGTAAAGCTCTGTCTTCAGGGACGATAATGACGGTACCTGAGGAGGAAGCCACGGCTAACT
ACGTGCCAGCAGCCGCGGTAATACGTAGGTGGCGAGCGTTGTCCGGATTTACTGGGCGTAAAGGGA
GCGTAGGCGGACTTTTAAGTGAGATGTGAAATACCCGGGCTCAACTTGGGTGCTGCATTTCAAACTG
GAAGTCTAGAGTGCAGGAGAGGAGAATGGAATTCCTAGTGTAGCGGTGAAATGCGTAGAGATTAGG
AAGAACACCAGTGGCGAAGGCGATTCTCTGGACTGTAACTGACGCTGAGGCTCGAAAGCGTGGGGA
GCAAACAGGATTAGATACCCGGGTAGT

Collinsella_ASV4245
CAGTGGGGAATCTTGCGCAATGGGGGGAACCCTGACGCAGCGACGCCGCGTGCGGGACGGAGGCC
TTCGGGTCGTAAACCGCTTTCAGCAGGGAAGAGTCAAGACTGTACCTGCAGAAGAAGCCCCGGCTAA
CTACGTGCCAGCAGCCGCGGTAATACGTAGGGGGCGAGCGTTATCCGGATTCATTGGGCGTAAAGC
GCGCGTAGGCGGCCCGGCAGGCCGGGGGTCGAAGCGGGGGGCTCAACCCCCCGAAGCCCCCGGAA
CCTCCGCGGCTTGGGTCCGGTAGGGGAGGGTGGAACACCCGGTGTAGCGGTGGAATGCGCAGATAT
CGGGTGGAACACCGGTGGCGAAGGCGGCCCTCTGGGCCGAGACCGACGCTGAGGCGCGAAAGCTG
GGGGAGCGAACAGGATTAGATACCCGGGTAG

Romboutsia_ASV9859
CAGTGGGGAATATTGCACAATGGGCGAAAGCCTGATGCAGCAACGCCGCGTGAGCGATGAAGGCCT
TCGGGTCGTAAAGCTCTGTCCTCAAGGAAGATAATGACGGTACTTGAGGAGGAAGCCCCGGCTAACT
ACGTGCCAGCAGCCGCGGTAATACGTAGGGGGCTAGCGTTATCCGGAATTACTGGGCGTAAAGGGT
GCGTAGGTGGTTTCTTAAGTCAGAGGTGAAAGGCTACGGCTCAACCGTAGTAAGCCTTTGAAACTGG
GAAACTTGAGTGCAGGAGAGGAGAGTGGAATTCCTAGTGTAGCGGTGAAATGCGTAGATATTAGGA
GGAACACCAGTTGCGAAGGCGGCTCTCTGGACTGTAACTGACACTGAGGCACGAAAGCGTGGGGAG
CAAACAGGATTAGATACCCGTGTAGTC
