## Supplemental Table 4 for "Adolescent autism and autoimmune diagnoses linked to infant gut bacteria whose prevalence is associated with at-risk genetics and/or diet": Supplementary_Table_4.pdf

**Supplemental Table 4. Permanova shows gut bacterial communities differ significantly by disorder groups.**

| | df | Sum of Squares | Mean Squares | F Model | $R^2$ | Pr(>F) | Sig. |
|---|---|---|---|---|---|---|---|
| *Binomial* | | | | | | | |
| Autoimmune vs. Autism vs. Control | 2 | 35914 | 17957 | 1.7197 | 0.01567 | 0.001 | *** |
| Autoimmune vs. Autism | 1 | 8796 | 8796.5 | 0.92107 | 0.01167 | 0.648 | |
| Autoimmune vs. Control | 1 | 25332 | 25332 | 2.3602 | 0.01208 | 0.001 | *** |
| Autism vs. Control | 1 | 13630 | 13630 | 1.295 | 0.00798 | 0.068 | .. |
| Disorder Development vs. Control | 1 | 27117 | 27117 | 2.5989 | 0.01183 | 0.001 | *** |
| *Bray-Curtis* | | | | | | | |
| Autoimmune vs. Autism vs. Control | 2 | 1.087 | 0.5434 | 1.4519 | 0.01326 | 0.009 | ** |
| Autoimmune vs. Autism | 1 | 0.4494 | 0.44936 | 1.2252 | 0.01547 | 0.138 | |
| Autoimmune vs. Control | 1 | 0.612 | 0.61228 | 1.6355 | 0.0084 | 0.014 | * |
| Autism vs. Control | 1 | 0.507 | 0.50716 | 1.3424 | 0.00827 | 0.063 | .. |
| Disorder Development vs. Control | 1 | 0.637 | 0.63745 | 1.7016 | 0.00778 | 0.009 | ** |
| *Jaccard (binary)* | | | | | | | |
| Autoimmune vs. Autism vs. Control | 2 | 1.066 | 0.53324 | 1.2494 | 0.01144 | 0.016 | * |
| Autoimmune vs. Autism | 1 | 0.465 | 0.46464 | 1.1014 | 0.01392 | 0.215 | |
| Autoimmune vs. Control | 1 | 0.582 | 0.58151 | 1.3621 | 0.00701 | 0.015 | * |
| Autism vs. Control | 1 | 0.509 | 0.50916 | 1.1867 | 0.00732 | 0.084 | .. |
| Disorder Development vs. Control | 1 | 0.602 | 0.60183 | 1.4095 | 0.00645 | 0.013 | * |

PERMANOVA results after testing for significant differences using three metrics: Binomial, Bray-Curtis, and Jaccard ($n = 219$).
Significance levels: '***' $p < 0.001$; '**' $p < 0.01$; '*' $p < 0.05$; '..' $p < .09$; '.' $p < 0.1$
