## Supplemental Table 5 for "Adolescent autism and autoimmune diagnoses linked to infant gut bacteria whose prevalence is associated with at-risk genetics and/or diet": Supplementary_Table_5.pdf

Table 3. Number of subjects with HLA Haplotype. Parentheses represent percentage of subjects. First number is number of subjects with at least one copy of the indicated haplotype. * indicates number of subjects with haplotype that are homozygous.

| HLA Haplotype | All Subjects (1748) | | Controls (1652) | | Cases (96) | | Celiac (24) | | Autism (24) | | Hypothy (4) | | Crohn's (5) | | JIA (9) | | T1D (12) | |
|---|---|---|---|---|---|---|---|---|---|---|---|---|---|---|---|---|---|---|
| DR4-DQ8 | 464(26.5) | *56(3.2) | 431(26.1) | *49(3.0) | 33(34.4) | *7(7.3) | 11(45.8) | *1(4.2) | 10(41.7) | *3(12.5) | 2(50.0) | - | 1(20.0) | - | 2(22.2) | - | 7(58.3) | *3(25.0) |
| DR3-DQ2 | 387(22.1) | *33(1.9) | 354(21.4) | *32(1.9) | 33(34.4) | *1(1.0) | 14(58.3) | *1(4.2) | 7(29.2) | - | 2(50.0) | - | 3(60.0) | - | 3(33.3) | - | 4(33.3) | - |
| DR15-DQ602 | 342(19.6) | *44(2.5) | 329(19.9) | *44(2.7) | 13(13.5) | - | 6(25.0) | - | 3(12.5) | - | - | - | 1(20.0) | - | 2(22.2) | - | 1(8.3) | - |
| DR1-DQ5 | 334(19.1) | *39(2.2) | 315(19.1) | *36(2.2) | 19(19.8) | *3(3.1) | 1(4.2) | - | 10(41.7) | *2(8.3) | 1(25.0) | - | - | - | 5(55.6) | *1(11.1) | 2(16.7) | - |
| DR5-DQ7 | 258(14.8) | *17(1.0) | 245(14.8) | *17(1.0) | 13(13.5) | - | 3(12.5) | - | 4(16.7) | - | 2(50.0) | - | 1(20.0) | - | 1(11.1) | - | 2(16.7) | - |
| DR7-DQ2 | 159(9.1) | *14(0.8) | 153(9.3) | *13(0.8) | 6(6.3) | *1(1.0) | 3(12.5) | - | 2(8.3) | *1(4.2) | - | - | - | - | - | - | 1(8.3) | - |
| DR13-DQ603 | 159(9.1) | - | 152(9.2) | - | 7(7.3) | - | 2(8.3) | - | 2(8.3) | - | - | - | 1(20.0) | - | 2(22.2) | - | - | - |
| DR4-DQ7 | 149(8.5) | *2(0.1) | 141(8.5) | *2(0.1) | 8(8.3) | - | 2(8.3) | - | 2(8.3) | - | 2(50.0) | - | 1(20.0) | - | 1(11.1) | - | - | - |
| DR8-DQ4 | 130(7.4) | *7(0.4) | 124(7.5) | *6(0.4) | 6(6.3) | *1(1.0) | 2(8.3) | - | 1(4.2) | - | - | - | - | - | 2(22.2) | - | 1(8.3) | - |
| D13-DQ604 | 117(6.7) | *5(0.3) | 112(6.8) | *5(0.3) | 5(5.2) | - | 2(8.3) | - | 1(4.2) | - | - | - | 1(20.0) | - | - | - | 1(8.3) | - |
| DR7-DQ9 | 88(5.0) | *7(0.4) | 86(5.2) | *7(0.4) | 2(2.1) | - | 1(4.2) | - | - | - | - | - | 1(20.0) | - | - | - | - | - |
| DR9-DQ9 | 56(3.2) | *6(0.3) | 53(3.2) | *6(0.4) | 3(3.1) | - | 1(4.2) | - | 1(4.2) | - | - | - | - | - | - | - | 1(8.3) | - |
| DR14-DQ503 | 14(0.8) | - | 12(0.7) | - | 2(2.1) | - | - | - | 1(4.2) | - | - | - | - | - | 1(11.1) | - | - | - |
| DR14-DQ5 | 7(0.4) | - | 6(0.4) | - | 1(1.0) | - | - | - | - | - | 1(25.0) | - | - | - | - | - | - | - |
| DR16-DQ502 | 5(0.3) | - | 5(0.3) | - | - | - | - | - | - | - | - | - | - | - | - | - | - | - |
| DR15-DQ601 | 3(0.2) | *1(0.1) | 3(0.2) | *1(0.1) | - | - | - | - | - | - | - | - | - | - | - | - | - | - |
| DR16-DQ5 | 2(0.1) | - | 2(0.1) | - | - | - | - | - | - | - | - | - | - | - | - | - | - | - |
| Undefined | 297(17.0) | | 283(17.1) | | 14(14.6) | | 4(16.7) | | 7(29.2) | | 1(25.0) | | 1(20.0) | | 0(0) | | 1(8.3) | |
